# Fentanyl initiation rate following the requirement for specialist approval

**DOI:** 10.1101/2024.03.13.24304188

**Authors:** Oren Miron, Yael Wolff-Sagy, Mark Levin, Esti Lubich, Jordan Lewinski, Maya Shpunt, Wiessam Abu Ahmad, Ilya Borochov, Doron Netzer, Gil Lavie

## Abstract

**Importance:** Healthcare organizations are exploring tools to address unwarranted fentanyl use which often leads to increased risk of addiction and overdose.

**Objective:** To assess the impact of a requirement for a specialist’s approval on fentanyl initiation for non-oncological pain.

**Design, Settings and Participants:** Retrospective cohort examination of fentanyl initiations and opioid dispensations for 4.4 million members of Clalit Health Services following a requirement for specialist’s approval for fentanyl initiation on July 2022, which was expanded 6 months later for continued use.

**Main Outcomes and Measures:** We analyzed the change in initiations of fentanyl in the year before and after the implementation and 95% confidence interval, with a sub-group analysis by age group. We also compared total opioid dispensation, fentanyl, and non-fentanyl in the 6th and 12th month after the implementation with the predicted rate based on pre-implementation rates.

**Results:** The fentanyl initiation rate in the year before the requirement was 711/1,000,000 capita, which decreased following the requirement by -81% (95% confidence interval:-77%; -85%). The decrease attenuated with age: at ages 0-17 years -100% (16%; -216%), at ages 18-39 years -88% (−78%; -97%), at ages 40-64 years -89% (−83%; -95%) and at ages 65 years and above -73% (−68%; -79%). In the 6th month after the requirement was implemented the morphine milligram equivalent from dispensation of total opioids and fentanyl was lower than predicted by 7% and 12% respectively, while non-fentanyl opioids dispensation was 3% higher than predicted. In the 12^th^ month after the initiation requirement, the dispensation of total opioids and fentanyl was lower than predicted by 26% and 39% respectively, while in non-fentanyl opioids it was 4% higher than predicted.

**Conclusions and Relevance:** Our results indicate that requiring specialist approval for fentanyl initiation for non-oncological chronic pain was associated with a decrease in fentanyl prescription initiations, especially among non-elderly patients. A decrease also occurred gradually in total opioid dispensations, further decreasing following the extension of the requirement to continuous fentanyl. These findings suggest that requiring specialist approval for non-oncological fentanyl initiations is likely an effective strategy to be considered by other healthcare providers.

**Question:** Was the requirement for specialist approval when initiating fentanyl for non-oncological pain followed by a decrease in fentanyl initiations and overall dispensing of opioids?

**Answer:** In this cohort study of 4.4 million members of Clalit Health Services without cancer, 81% decrease in fentanyl initiations was observed in the year following the implementation of the requirement for specialist approval. After 6 months from implementation, dispensed morphine milligram equivalent from opioids decreased by 7%.

**Meaning:** These findings suggest that requiring specialist approval for fentanyl initiation in non-oncological pain can help reduce fentanyl initiations and overall opioids dispensed and is likely an effective strategy to be considered by other healthcare providers.

## Introduction

Studies have shown that fentanyl initiations for non-oncological chronic pain are often given despite contra-indications, such as not proving prior opioid tolerance for weaker opioids.^1^ For example, a study has shown that around 50% of fentanyl initiations in Canada occurred without sufficient opioid tolerance.^2^ It should be noted that examining contraindications for fentanyl can require considerable time and knowledge. For example, identifying prior opioid tolerance requires collecting records on all prior opioid consumption by type and dosage, converting each type to morphine milligram equivalent (MME), and summing the daily dose for each of the last 7 days, to ensure they passed 60 morphine milligram equivalent per day.^2^

Initiation of fentanyl without prior tolerance to weaker opioids during the past 7 days was associated with respiratory arrest, and fentanyl prescription is associated with opioid use disorder and with opioid overdoses causing severe respiratory depression.^3–5^ In recent years, opioid overdose rates have increased in many developed countries, with the United States suffering from 80,000 overdose deaths in 2021.^6^

Dispensing of fentanyl has significantly increased in Israel over the past decade, especially among non-oncological younger patients.^7^ This led Israel to a 120% increase in opioid consumption between 2011 and 2016, which was the largest increase in the OECD and twice as high as that of the United Kingdom which had the 2^nd^ largest increase.^8^ This increase in fentanyl initiations among non-elderly and low socio-economic Israelis was followed by an increase in fentanyl abuse, dependence, and overdose.^5,9^

These concerning patterns led Clalit Health Services (Clalit), the largest healthcare organization in Israel, to require a specialist’s approval for the initiation of fentanyl for non-oncological chronic pain, which was implemented on July 20, 2022. The approval requirement was extended to continuous fentanyl use on January 29, 2023. Our objective was to assess the impact of the specialist requirement for fentanyl initiation, and its long-term effect on fentanyl and non-fentanyl opioid dispensation.

## Methods

### Study Design

A retrospective cohort study among members of Clalit was conducted from July 20, 2021, to July 19, 2023. In order to identify initiations of fentanyl, prior prescriptions were screened starting from January 2013 when prescription data became widely available in Clalit.

### Study Population

Members of Clalit, which insures an estimated 52% of Israel’s population. Patients with an oncological diagnosis recorded in the Electronic Health Record database of Clalit were excluded from the study since they were not affected by the requirement for specialist approval for fentanyl invitation.

### Data Sources and organization

Data was queried from the Electronic Health Record database of Clalit, which pools the data from its operational systems to a data warehouse used for research and healthcare policy queries. The following demographic data were extracted for each participant: age, sex, socioeconomic status (SES), and sector (general Jewish population, Ultraorthodox Jewish population, and Arab population). Socioeconomic status and sector were defined based on the place of residence and analyzed by Small Statistical Areas (SSA). SSAs in Israel contain 3,000-4,000 people and are defined by the Central Bureau of Statistics of Israel.^10^ The prescribing data was extracted for fentanyl, which has the Anatomical Therapeutic Chemical (ATC) code of N02AB03. The fast-release oral fentanyl is only approved for oncological pain in Israel and was therefore excluded. For the analysis of the other opioids, data was extracted for the other drugs under the opioids ATC (N02A), which were Buprenorphine (N02AE01), Tramadol (N02AX02, N02AJ13), Oxycodone (N02AA55, N02AJ17, N02AA05), Codeine (N02AJ09, N02AJ06), Morphine (N02AA01), Pethidine (N02AB02), and Propoxyphene (N02AC54).

### Statistical Analysis

The rate of fentanyl prescription initiation per 1,000,000 Clalit members was calculated in the year before fentanyl initiations required specialist approval (July 20, 2021 to July 19, 2022), and compared to the year following the requirement (July 20, 2022 to July 19, 2023). The rate difference, with its 95% confidence interval and percent change was calculated. A sub-group analysis repeated this comparison by age (0-17, 18-39, 40-64, and 65 years or older).

An additional analysis examined the dispensing of all opioids, fentanyl, and non-fentanyl opioids based on morphine milligram equivalent (MME) in the 6^th^ and 12^th^ month after the initiation requirement started. This secondary analysis utilized dispensations rather than prescriptions, as this measure is more accurate in evaluating the extent of actual use in MME.

This mean difference was compared to the predicted monthly rate based on linear regression of the monthly average rate before the requirement for specialist approval for fentanyl initiation. All analyses were performed using R-statistical software.

### Ethical considerations

The study was approved by the Institutional Review Board of Clalit Health Services.

## Results

In the year before requiring specialist approval for prescribing fentanyl to non-oncological patients (20 July 2021 to 19 July 2022), Clalit had 4.8 million members. Of those, 320,197 were excluded from the study due to a previous oncological diagnosis and 16,975 were excluded from the initiation analysis due to a previous prescription of fentanyl. Therefore, the initiation analysis included 4.417 million members in the first year, while in the following year (20 July 2022 to 19 July 2023) it included almost 4.5 million members (Table 1).

**Table 1:**
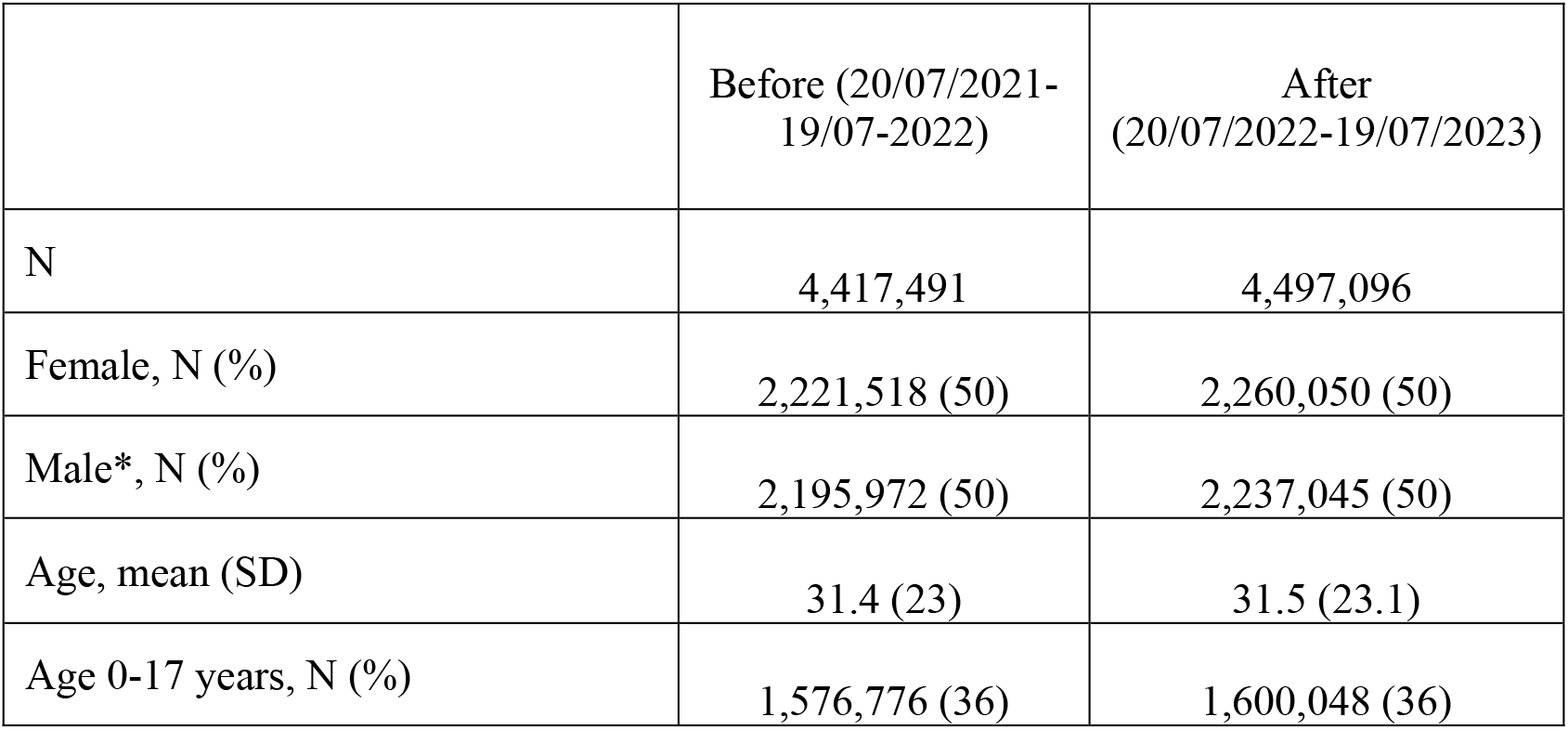
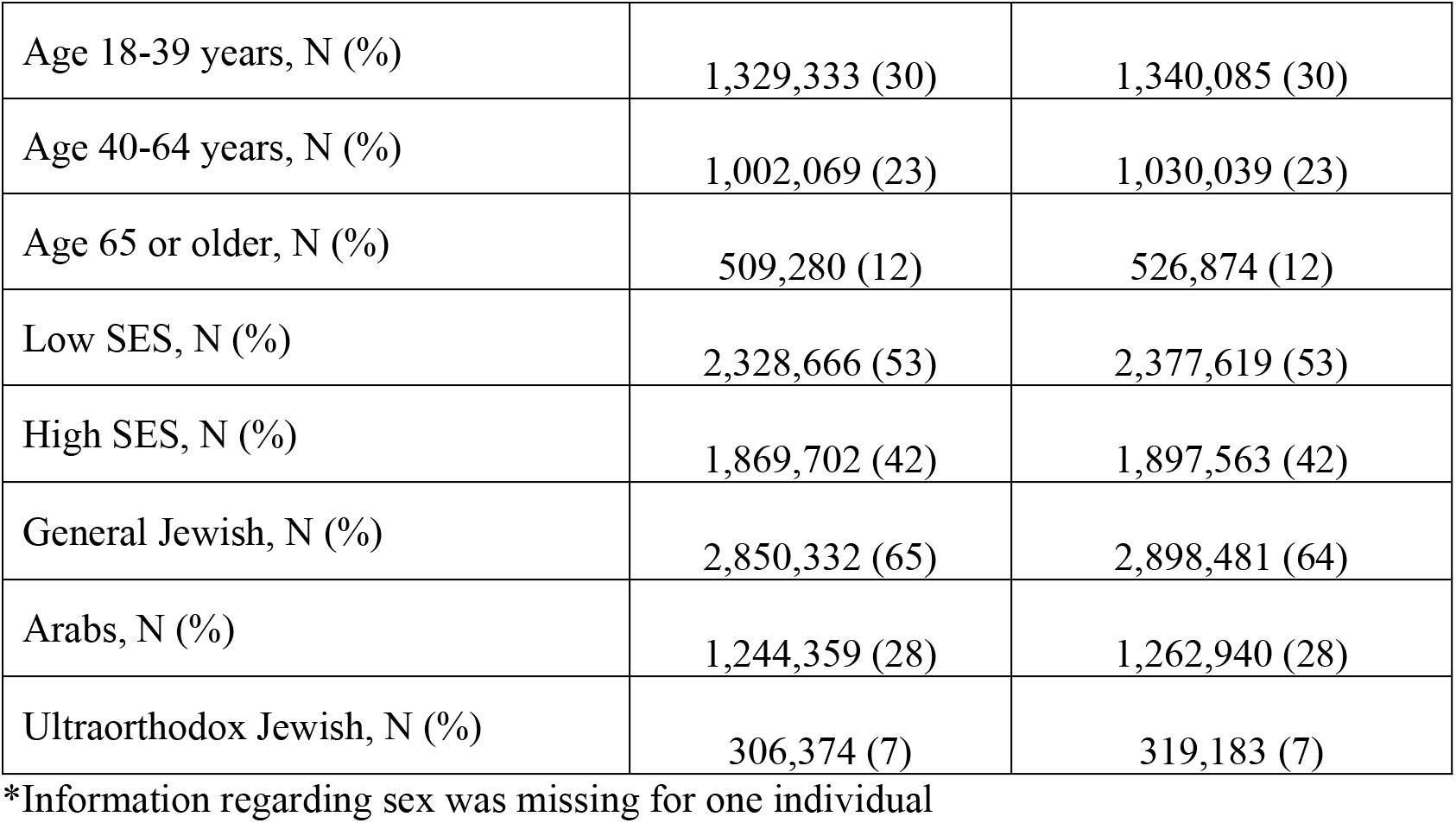
Study population characteristics.

|  | Before (20/07/2021-19/07-2022) | After (20/07/2022-19/07/2023) |
| --- | --- | --- |
| N | 4,417,491 | 4,497,096 |
| Female, N (%) | 2,221,518 (50) | 2,260,050 (50) |
| Male*, N (%) | 2,195,972 (50) | 2,237,045 (50) |
| Age, mean (SD) | 31.4 (23) | 31.5 (23.1) |
| Age 0-17 years, N (%) | 1,576,776 (36) | 1,600,048 (36) |
| Age 18-39 years, N (%) | 1,329,333 (30) | 1,340,085 (30) |
| Age 40-64 years, N (%) | 1,002,069 (23) | 1,030,039 (23) |
| Age 65 or older, N (%) | 509,280 (12) | 526,874 (12) |
| Low SES, N (%) | 2,328,666 (53) | 2,377,619 (53) |
| High SES, N (%) | 1,869,702 (42) | 1,897,563 (42) |
| General Jewish, N (%) | 2,850,332 (65) | 2,898,481 (64) |
| Arabs, N (%) | 1,244,359 (28) | 1,262,940 (28) |
| Ultraorthodox Jewish, N (%) | 306,374 (7) | 319,183 (7) |

\*Information regarding sex was missing for one individual

In the year before requiring specialist approval for prescribing fentanyl to non-oncological patients, the initiation rate was constant with 708 annual initiations per 1,000,000 capita. Following the requirement, the prescription initiation rate decreased by 81% throughout the following year (Table 2-annual rates; Figure 1-daily rates).

**Table 2:** Fentanyl prescription initiations per year.

|  | Fentanyl initiations per year per 1,000,000 capita |  |  | % Change (95% CI) |
| --- | --- | --- | --- | --- |
|  | Rate Before*<br>(20/07/2021-19/07-2022) | Rate After**<br>(20/07/2022-19/07/2023) | Rate difference<br>(95% CI) |  |
| All | 710.8 | 135.9 | -547.9; -602 | -81% (-77.1; -84.7) |
| Age 0-17 years | 1.9 | 0 | 0.3; -4.1 | -100% (15.8; -215.8) |
| Age 18-39 years | 381.4 | 47.0 | -299.2; -369.5 | -88% (-78.4; -96.9) |
| Age 40-64 years | 1087.7 | 116.5 | -903.4; -1039.1 | -89% (-83.1; -95.5) |
| Age 65 or older | 3023.9 | 812.3 | -2042.2; -2380.8 | -73% (-67.5; -78.7) |

\* Started fentanyl during the year before the intervention
\*\* Started fentanyl during the year after the intervention

**Figure 1:**
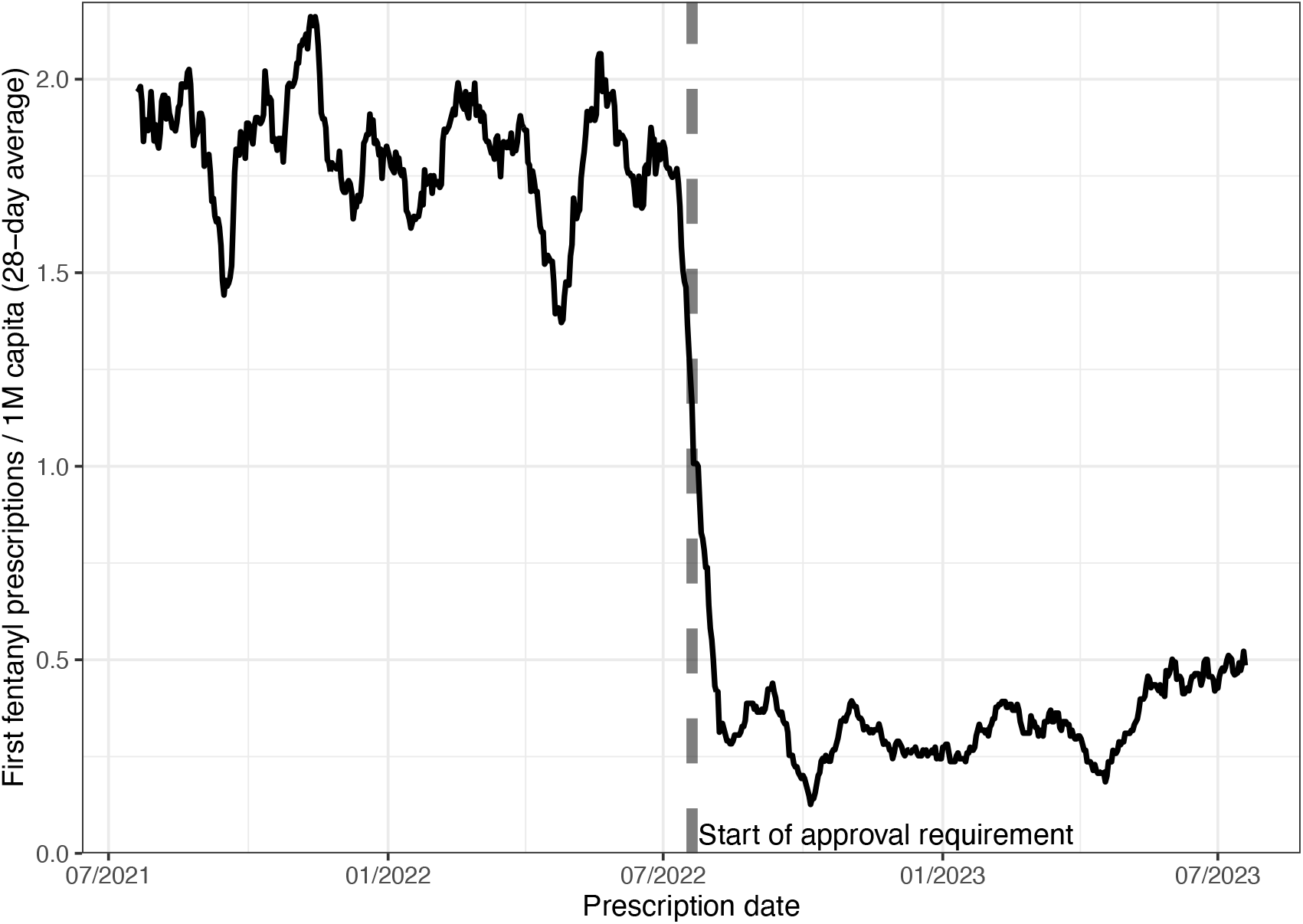
Fentanyl prescription initiation before and after requiring approval. Y-axis indicates fentanyl prescription. initiations per 1,000,000 members of Clalit Health Services with a 28-day moving average. X-axis indicates the date of the fentanyl prescription. The dotted line indicated the date when fentanyl initiation started to require specialist approval.

**Figure 2:**
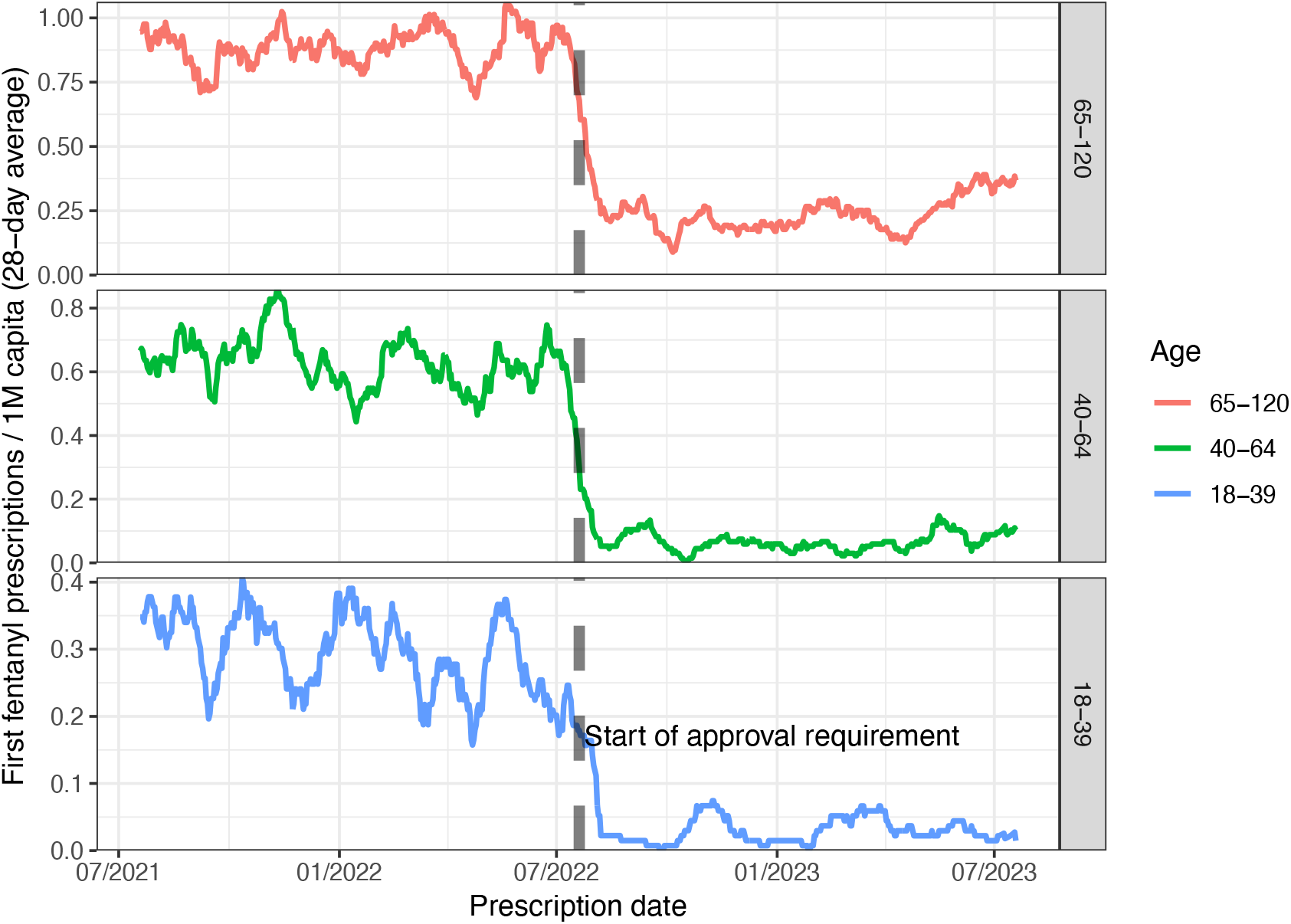
Fentanyl prescription initiation before and after requiring approval by age. Y-axis indicates fentanyl prescription initiations per 1,000,000 members of Clalit Health Services with a 28-day moving average. X-axis indicates the date of fentanyl prescription. The dotted line indicated the date when the requirement for a specialist’s approval for fentanyl initiation was implemented. The panels from top to bottom are ages 65 years and above, ages 40-64 years, and ages 18-39 years. Ages 0-17 are not shown as there were not enough initiations to display on a daily rate.

In the year before requiring a specialist approval for non-oncological fentanyl, the daily prescription initiation rates per 1,000,000 capita at ages 0-17 years, 18-39 years, 40-64 years, and 65 years and above were 2, 381, 1080, and 2980 respectively. In the following year, the yearly rate of fentanyl prescription initiations decreased at ages 0-17 years by -100% (−216%; 16%), at ages 18-39 years by -88% (−97%; -78%), at ages 40-64 years by -89% (−95%; -83%) and at ages 65 years and above by -73% (−79%; - 68%) (Table 2).

In the 6^th^ month after starting the requirement for specialist approval, the MME per capita dispensed from all opioids was 18.24, which was 7% less than the predicted 19.67 (95%-CI 19.55; 19.79). The MME per capita dispensed from fentanyl was 12.01, which was 12% less than the predicted 13.6 (95%-CI 13.52; 13.68). The MME per capita dispensed from non-fentanyl opioids was 6.23, which was 3% more than the predicted 6.07 (95%-CI 6.02; 6.12).

At the 12^th^ month after starting the requirement for specialist approval for initial use, which was the 6th month after requiring approval for continued use, the MME per capita dispensed from all opioids was 15.16, which was 26% less than the predicted 20.55 (95%-CI 20.37; 20.73). The MME per capita dispensed from fentanyl was 8.73, which was 39% less than the predicted 14.37 (95%-CI 14.25; 14.49). The MME per capita dispensed from non-fentanyl opioids was 6.43, which was 4% more than the predicted 6.18 (95%-CI 6.11; 6.26, Figure 3).

**Figure 3:**
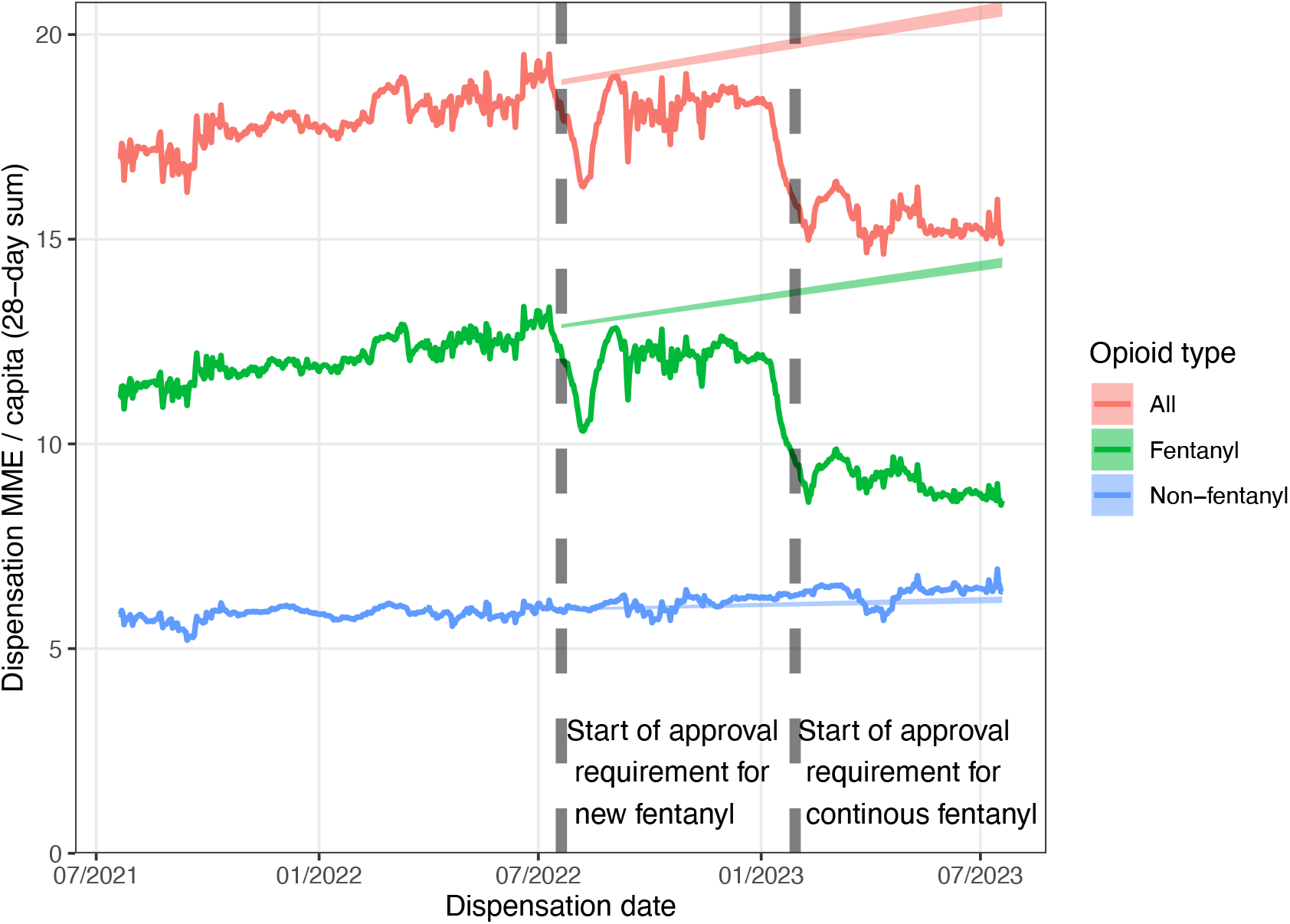
Overall morphine milligram equivalent (MME) from opioid dispensation. Y-axis indicates fentanyl dispensations morphine milligram equivilent (MME) per member of Clalit Health Services with a 28-day moving average. X-axis indicates the date of fentanyl prescription. The left dotted line indicates the date when the requirement for specialist’s approval upon fentanyl initiation was implemented, and the right indicates the date when the requirement for specialist’s approval was extended to continued fentanyl roval. Red indicates all opioids, green indicates fentanyl, and blue indicates non-fentanyl opioids. The shaded area indicates the 95% confidence interval of the predicted rate based on linear regression on the rate until the start of the fentanyl limit.

## Discussion

In this cohort study of Clalit Health Service patients, implementing a requirement for specialist approval for fentanyl initiation for non-oncological patients was followed by a sharp decrease in fentanyl prescription initiations, which remained for at least a year. The requirement was also followed by a gradual decrease in MME from dispensing fentanyl and overall opioids and a small gradual increase in MME from dispensing non-fentanyl opioids.

The largest fentanyl prescription initiation decrease occurred among non-elderly members, which could stem from non-elderly members having higher rates of fentanyl prescribing that the specialists considered unwarranted. Such unwarranted fentanyl prescribing among non-elderly members could relate to reports of increased fentanyl dispensing and abuse in recent years.^5,7^ These reports have also raised awareness of the risks of fentanyl initiation, especially at younger ages, which in turn could have contributed to the decrease in fentanyl initiation at younger ages.

The requirement for specialist approval for fentanyl initiation was followed by a decrease in MME from fentanyl dispensation and from any opioid dispensations, despite a smaller compensatory increase in non-fentanyl dispensation. This could stem from fentanyl being more potent and addictive compared to most prescription opioids, which were less likely to lead to an increase in dispensed MME. Another possible reason could be that codeine, morphine, and oxycodone have a daily milligram limit before requiring approval according to Israeli legislation.^10^ However, such approval does not apply to tramadol and buprenorphine.

Israel limited the initiations of the weak opioid propoxyphene in 2011 due to its adverse effects, and the limitation was followed by increased dispensing and abuse of stronger opioids.^7,11^ This example, together with our finding, suggests that when limiting the prescribing of an opioid, it is necessary to ensure it would not be replaced by more potent and abuse-prone opioids.

The requirement for specialist approval for fentanyl prescribing in July 2022 affected new fentanyl users, who otherwise would have probably started with low fentanyl doses and gradually increased them. This could explain the gradual decrease in MME dispensation following the requirement from July 2022 to January 2023. The MME form opioid dispensation decreased more sharply after January 2023, when the requirement for specialist approval was extended to continuous fentanyl users. The continuous users often use higher daily MME than new users, which makes the limitation on their fentanyl dispensation have a faster effect on the total MME dispensed. It is worth noting that the specialist approval was given to continuous fentanyl users in most cases, to prevent an abrupt discontinuation of fentanyl use, which could have caused insufficient pain relief and strong withdrawal symptoms. The risk of such a scenario was shown in the U.S. when the abrupt discontinuation of abuse-prone OxyContin in 2010 led many users to abuse heroin.^12^

The impact of previous opioid prescribing policy changes was examined by McGinty et al, which evaluated the impact of thirteen treatment states that implemented different opioid prescribing laws, and found mostly small-magnitude, non-significant changes.^13^ However, a more recent analysis of setting-specific limits on prescription showed significant decreases in initial prescribing length following the implementation of the law.^14^ The requirement of specialist approval allows for an even more tailored decision since the specialist takes into account the patient’s prescribing history and abuse risk. This process increases accuracy but also time, which could be shortened in the future with an algorithm that summarizes the relevant data for the specialist and the clinician who first prescribed the fentanyl. For example, such an algorithm could calculate if the patient has medications or conditions that are counter-indicated for fentanyl.

Our main limitation is that our data is based on digital prescriptions and does not include hand-drawn prescriptions. Hand-drawn prescriptions are not as common as digital prescriptions but could hold special importance as they are harder to track and easier to forge, which led to proposals to limit their use for prescribing fentanyl.^15^

Another limitation is that the low rate of fentanyl initiation in minors prevented us from determining the effect of the requirement despite the sharp decrease and the large number of participants. The low rate of fentanyl prescriptions in minors for non-oncological pain likely stems from the extra caution taken when prescribing fentanyl to this unique population. Moreover, there have been reports of minors purchasing prescription fentanyl from adults and abusing it, which suggests that limiting unwarranted fentanyl prescriptions to adults could help reduce its abuse among minors.^9^

The decision to exclude cancer patients from requiring a specialist’s approval stems from their increased need for pain relief and from the increased medical monitoring that they are given, which helps prevent unwarranted use and detect addiction. Future policy makers could consider expanding this exclusion criteria for other severe chronic and terminal conditions, and cloud also consider not excluding specific types of cancer that are much less severe, such as early-stage basal cell carcinoma.^16^

In conclusion, requiring specialist approval for fentanyl initiation for non-cancer chronic pain was followed by a sharp decrease in fentanyl initiations and a gradual decrease in the total MME dispensed from opioids. A similar requirement might benefit other countries with unwarranted initiations of fentanyl or other strong opioids, especially if safer alternatives are given to patients to prevent them from resorting to dangerous drugs.

## Data Availability

The Institutional Review Board approval does not allow the release of individual patient data

